# Utilization profile and diagnostic completeness in a hospital emergency service in Medio Paraiba Fluminense, Brazil: an observational study from 2023 to 2025

**DOI:** 10.64898/2026.11.26.26368978

**Authors:** Alexandre Rodrigues de Souza Soares, Tamyres Silva Salles Cardoso, Yago Aragão Lessa, Luís Gustavo Ferreira de Souza, João Victor Cordeiro da Silva, Rachel Rocha Ferreira, André Manoel Correia dos Santos, Aluana Santana Carlos, André Costa Ferreira

## Abstract

High demand for low-complexity care in emergency services reflects structural and operational failures in Primary Health Care, incomplete medical records. The article measures the impact of care overload on the rate of incompleteness of medical records in a tertiary reference hospital complex. It is a single-center, observational, descriptive and analytical study of emergency visits performed between 2023 and 2025 at Santa Casa de Misericórdia in Barra Mansa, RJ, using the Manchester protocol, geoprocessing and descriptive/analytical statistics. Of the total of 175,004 admissions, low-complexity cases represented 67.85% of the demand and georeferencing demonstrated a strong concentration of Basic Health Units in the immediate surroundings of the hospital, aggravated by a care void during extended hours, and there was an alarming rate of medical record incompleteness of 24.33%, concentrated in lower-severity visits. The work demonstrates that saturation by low-complexity demands and the high rate of incomplete records are a direct reflection of the inversion of the care flow. This overload degrades the quality of health information, creating critical gaps for epidemiological surveillance and public management.

## INTRODUCTION

The organization of urgency and emergency services in hospital units constitutes one of the greatest challenges for the consolidation of the Unified Health System (SUS) in Brazil, given that they are configured as a strategic component of the Health Care Networks (RAS), as they concentrate care for various acute conditions that require rapid assessments and interventions of medium to high complexity^1^. Furthermore, it is necessary to highlight that urgency and emergency services are part of a set of health care categories, which, according to current RAS stratifications, advocate for an integrated, decentralized, and regionalized model, in which Primary Health Care (PHC) should act as the coordinating center of care and the main entry point of the SUS. However, the “diversion” of the population from PHC to urgency and emergency services, with the consequent persistence of high demand and saturation in high-complexity hospital services, signals structural restrictions in the articulation of the care network^2^.

The immense demand for hospital urgency units for the resolution of less severe health problems is multifactorial and significantly linked to social, cultural, geographical, and organizational factors. A large part of the population remains with the social and cultural perception focused on the old model of centralization of health care, since it credits greater efficacy to these hospital sectors due to greater technological availability and the possibility of immediate therapy. Corroborating this, organizational barriers in PHC, such as limitations in absorbing spontaneous demands, rigid schedules, and challenges in immediate resolution, impel the user to seek the emergency room as an operational shortcut to guarantee care^3,4^.

A well-established way to resolve this problem is the use of the Manchester Triage System (MTS), which aims to classify risk and order waiting time, prioritizing care according to the patient’s clinical severity. Furthermore, the very use of the MTS ends up revealing the expressive demand composed of users with lower severity, and how the large volume of potentially outpatient patients influences both the time and the quality of care for medium and high complexity patients^6,7^.

This distortion in the care flow generates significant impacts on clinical efficiency and patient safety, in addition to generating a disproportionate allocation of resources and professionals for the management of conditions capable of being monitored in primary care, compromising the hospital management capacity for clinically more serious events This operational overload is associated with determinants that can increase fragility of the health surveillance and public management system, one of the most critical being the failures in medical record entries such as lack of specification of the International Statistical Classification of Diseases and Related Health Problems (ICD-10), which increases incompleteness, hinders health management planning and compromises the integrity of databases essential for epidemiological monitoring.

This issue is of extreme relevance since these records are fundamental for feeding national databases such as SINAN (Information System for Notifiable Diseases) and SIH/SUS (Hospital Information System), and the incompleteness of these entries directly affects the quality of these databases, which impacts public health administration since these fragmented and/or incomplete data prevent managers from precisely identifying priorities for interventions and resource investment, perpetuating the fragility of the care system as well as intensifying regional inequalities in the provision of medium and high complexity public services^7,8,9^.

Although several studies focused on collective health address the phenomenon of “overcrowding” and its consequent partial obstruction in emergency departments and this serious problem of incomplete medical records, the persistence of large gaps in public health knowledge that simultaneously address the relationship between long-term care data and geospatial data analysis of urban grids in relation to the distribution of health units and tertiary reference centers, as well as the measurement of the impact of incomplete medical records on public management strategies.

Given this, the present study describes the potential determinants of care saturation in an important regional health hub, evaluating the temporal and spatial distribution of primary care with the overload of tertiary care, in order to support new reorganization strategies for the Health Care Networks (RAS), and thereby describe these systemic and structural determinants of emergency sector care saturation using a tertiary hospital complex of important regional reference, located in the municipality of Barra Mansa (RJ).

## METHODOLOGY

### Study design and investigation period

The present work is a single-center, observational, descriptive, and analytical study. The investigation evaluated the urgent and emergency service of the tertiary hospital complex of the Santa Casa de Misericordia of Barra Mansa, located in the State of Rio de Janeiro. The period of sampling and analysis took place during the triennium from January 2023 to December 2025.

### Data collection and care flow

The data were extracted from the care information system of the reception and the electronic hospital medical record. The observed care routine follows the following sequence: reception and registration -clinical triage (Manchester Triage System protocol, adapted according to the guidelines of the Ministry of Health^10^ (red/emergency, yellow/urgent, green/less urgent and blue/non-urgent) -Clinical referral (direction to the necessary sector, according to the severity and the care outcome and the diagnostic coding based on ICD-10)^11^.

### Sample selection and STROBE guidelines

The method of selecting the samples followed the international recommendations of the Strengthening the Reporting of Observational Studies in Epidemiology (STROBE) structured in three stages: Total population admitted (N = 175.004): Primary admission records made at the hospital reception during the triennium. Sample showing completeness of medical record filling in the hospital profile (n = 132.418). Sample showing incompleteness of medical records (n = 42.586) due to the absence of the ICD-10 code record.

### Information Quality Indicators

The evaluation of the quality of correct and complete filling of epidemiological data was measured by two indicators: medical record completeness rates (CR), proportion of records with ICD-10 code that were duly filled out through^12^, the formula CR = (n/N) x 100, where n represents the sample with completeness of ICD-10 completion and N represents the total admitted population) and proportion of incompleteness (P_inc_), percentage of records without ICD-10 code specification, through the formula (P_inc_ = 100% -TC = 100% -75,67% = 24,33%).

### Spatial and geographic accessibility analysis

For the spatial evaluation of health services, analyses were performed of georeferencing of Basic Health Units (UBS) and the hospital establishment of reference of the municipality of Barra Mansa, RJ. The territorial mesh and municipal limits were obtained from the official cartographic base provided by the Brazilian Institute of Geography and Statistics (IBGE)^13^. Data from the national registry of health establishments (CNES) were also used to evaluate the quantity, location an functioning of the basic health units in the municipality of Barra Mansa, RJ^14^.

The processing of geographic data, the creation of thematic maps and spatial calculations were performed with the aid of the free software QGIS. From the geographic coordinates, the Euclidean distance lines between each basic unit and the reference hospital were calculated, stratified by metric proximity ranges. In order to identify patterns of concentration and territorial dispersion of the care network, a weighted Kernel density map was generated.

### Study limitations

As this is a study with a unicentric design, it is not perfectly generalizable to the entire state of RJ, and the use of secondary administrative data makes it impossible to establish a clear causal relationship, limiting the study to a descriptive and analytical approach.

### Statistical processing and analysis

The data were subjected to descriptive and longitudinal statistical analysis. Continuous variables were expressed as annual means and 95% Confidence Interval (Mean and 95% CI). Categorical variables (complexity levels, risk classification colors, care outcomes and ICD-10 records) were presented in absolute frequencies (n) and relative (%).

To evaluate the temporal evolution and the care profile throughout the triennium, a two-way analysis of variance (Two-way ANOVA) with Tukey’s multiple comparisons test was used. The analysis of the distribution of care outcomes was conducted by means of the Chi-Square test. The data were organized, tabulated, and structured in an electronic database with the aid of the Microsoft Excel software, in which the consistency verification and duplicate debugging were performed. Statistical analyses and graphic elaboration were conducted in the GraphPad Prism software, and georeferencing analyses and thematic cartography were performed in the free software QGIS, adopting a level of significance of 5% (p < 0.05).

### Ethical aspects

The study was conducted in accordance with the ethical precepts established by Resolution No. 466/2012 of the National Health Council (CNS)^13^. The research project was submitted and approved by the Research Ethics Committee (CEP), under the Certificate of Presentation of Ethical Appreciation (CAAE) No. 92484025.4.0000.8044 (Approval Opinion No. 7.928.769). As the research used exclusively secondary, unidentified data, extracted from the institutional database of the region’s reference hospital, with no direct contact with patients and no risk of identification, a waiver of the Informed Consent Form (ICF) was requested, ensuring the secrecy and confidentiality of the information.

### Declaration of artificial intelligence (AI) use

In compliance with editorial guidelines and orientations, the authors declare that the use of a Generative Artificial Intelligence tool (ChatGPT / Large Language Model -LLM) was exclusively for technical support in academic writing assistance. It was used in a supervised manner in the stages of: (a) assistance in formal adaptation of formatting and adjustment of the strict character count in the Abstract, Abstract sections; (b) support in the translation and spelling/grammatical review process of the abstracts into English; and (c) support in the textual and stylistic organization of the manuscript. We emphasize that the resource was not used for generation, alteration, analysis, or interpretation of the primary research data, nor in the formulation of the study’s conclusions. All sections were duly prepared, reviewed, and validated, and the authors assume full responsibility for the intellectual content and the integrity of the final version of the manuscript.

## RESULTS

### Characterization of emergency care and clinical outcomes in the triennium

The evaluated triennium (2023–2025) recorded a total of 175,004 visits in the hospital’s emergency department, showing a progressive growth trend in the annual volume of records, which went from 51,643 in 2023 to 62,199 in 2025. Regarding risk stratification, there was a significant and persistent predominance of visits classified as low complexity (green and blue), which together represented a demand that varied from 69.64% to 65.12%, concentrating the majority of admitted cases. In contrast, cases classified as yellow (urgent) showed a gradual increase from 29.63% to 34.25% over the period, while red cases (emergencies) remained stable and with lower percentage representation, fluctuating between 0.54% and 0.73%. Regarding clinical outcomes, medical discharge represented the majority conduct of the care flow, stabilized at around 76.62% of the total for the triennium, while hospital admissions corresponded to 15.29% of general outcomes, and transfers and deaths maintained residual and constant rates, recording respectively 0.20% and 0.16% of the total records. Finally, the assessment of record completeness showed a progressive improvement in the filling of medical records and care forms, whose complete data rose from 70.92% in 2023 to 83.94% in 2025, though still with significant percentages (Supplementary Table 1).

### Sample Selection Dynamics and Record Quality

During the 2023 -2025 triennium, the emergency service of Santa Casa de Misericordia de Barra Mansa recorded a volume of 175,004 admissions at the emergency care reception submitted to reception with risk classification by the Manchester protocol. Of this eligible population, 42,586 medical records showed an absence of diagnostic coding (ICD-10), configuring a Completeness Rate of 75.67% (n = 132.418) for hospital analysis and an incompleteness rate of 24.33% of records null or unspecified in the database, which were summarized in the flow diagram (Figure S1).

### Prevalence of low-complexity demand in the hospital emergency sector

The evaluation of the care profile during the triennium (2023 – 2025) demonstrates a marked disproportion in care demand at the hospital unit, with care for low complexity (green and blue classification) corresponding to 67,85% (n = 118.737 / *p = 000.1*), while high-complexity demands (yellow and red classification) were responsible for 32,15% (n = 56.267) of cases (Figure 1).

**Figure 1.**
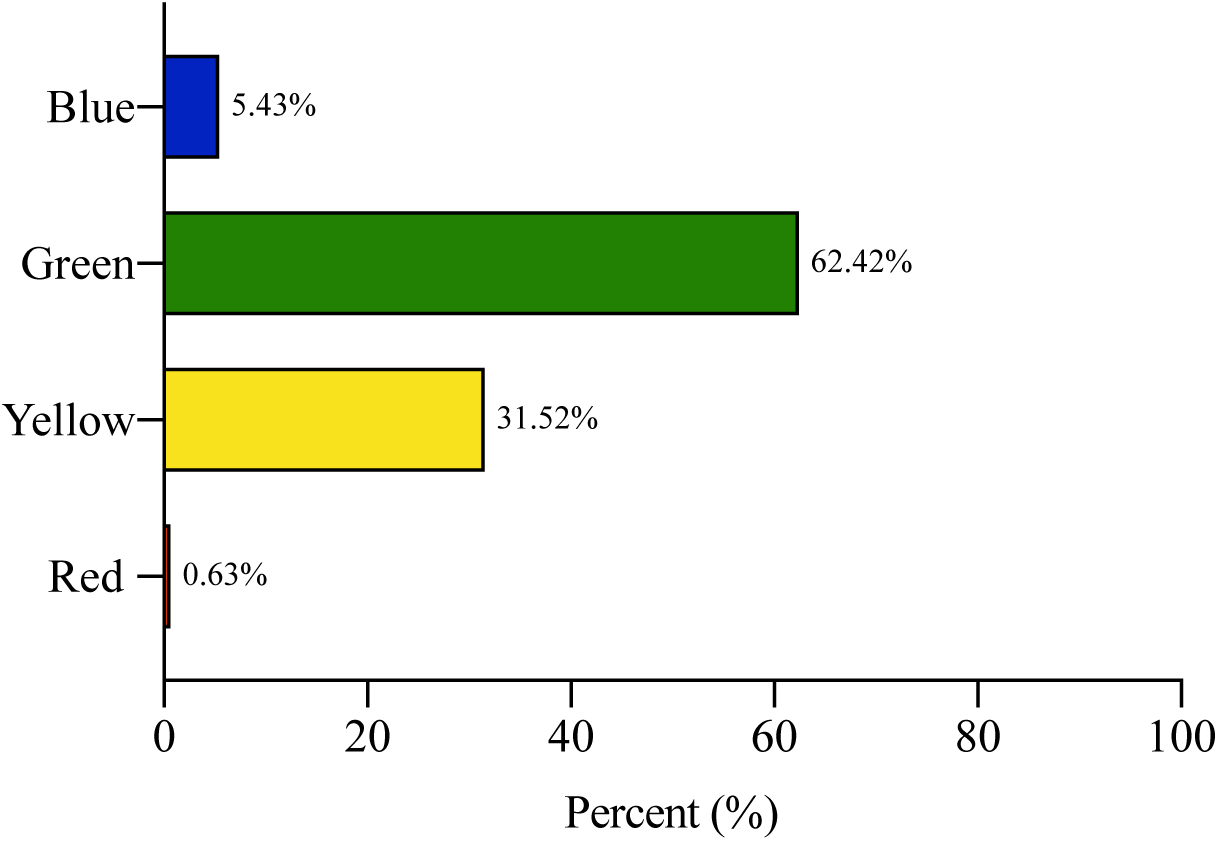
Distribution of emergency care according to annual volume, risk classification by the Manchester protocol, care outcomes, and completeness of records in the 2023 to 2025 triennium. Percentage distribution by categories (blue, green, yellow, red), where there was a significant difference between the low-complexity risk categories (blue and green) in high-complexity categories (yellow and red) by the Chi-Square test (X^2^ = 132.849, df = 1, *p < 0,0001*).

### Profile of care and distribution of care outcomes in the hospital emergency department in the 2023–2025 triennium

Regarding the profile of care outcomes (Figure 2A), the distribution of closures showed a higher proportion of discharges post-care and medication, revealing a predominance of outpatient resolution in the open-door sector itself and corroborating the values of the analyses of the low-complexity categories. The highest average annual of outcomes consisted of discharge after medical consultation (n = 25.977 cases/year) and discharge after administration of medication/direct observation (n = 18.521,33 cases/year). Hospital admissions represented a significant fraction of (n = 8.954,66 cases/year), followed by dropout rates (n = 4.514 cases/year) and occasional occurrences of deaths and other outcomes.

**Figure 2.**
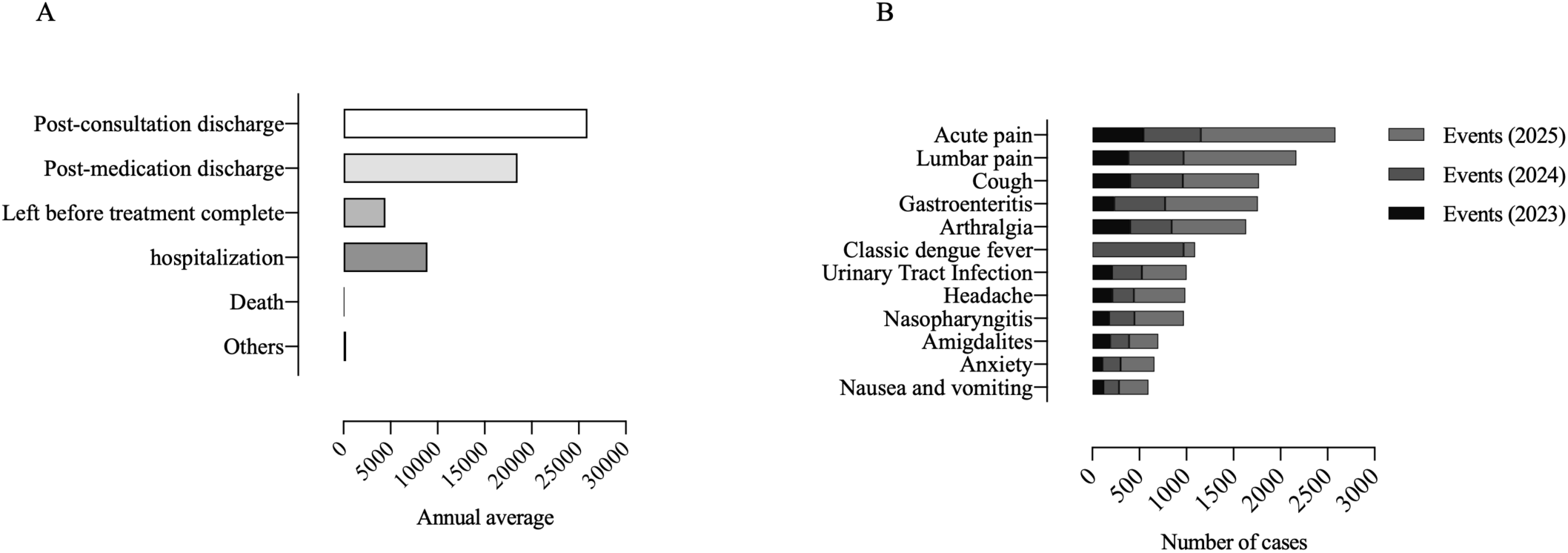
Distribution of care outcomes at the emergency service of Santa Casa de Misericordia, Barra Mansa, RJ (2023 – 2025). (A) Annual average by type of care outcome, showing a statistically heterogeneous and significant predominance of hospital discharges in relation to other outcomes by the Chi-Square test (*p < 0.05*). (B) Absolute number of cases of the main occurrences recorded per year (2023–2025). Source: Prepared by the authors based on hospital care files. Note. UTI – urinary tract infection.

Among the medical records validated by ICD-10 code (Figure 2B), the main clinical occurrences recorded in the triennium were concentrated in cases of acute pain, low back pain/lumbago, cough, diarrhea and infectious gastroenteritis, and joint pain/arthralgia.

### Temporal trend of care profile and distribution of care outcomes in the hospital emergency department in the 2023–2025 triennium

The longitudinal analysis of the triennium (2023–2025) demonstrated a significant difference between low-complexity care (green with an average of around 3,034 visits/month) and high-complexity care (yellow with an average of around 1.532 visits/month and red with an average of around 30 visits/month), both in the analyses between the different categories as well as in relation to the annual progression, highlighting a stability in the annual average of visits for the lower severity categories (Figure 3A). These visits corresponded to a proportion greater than two visits of low complexity for each one of high complexity in the years 2023 to 2024 and despite showing a drop in proportion in 2025, this reduction was not statistically significant, maintaining a pattern of predominance in low complexity visits (Figure 3B).

**Figure 3.**
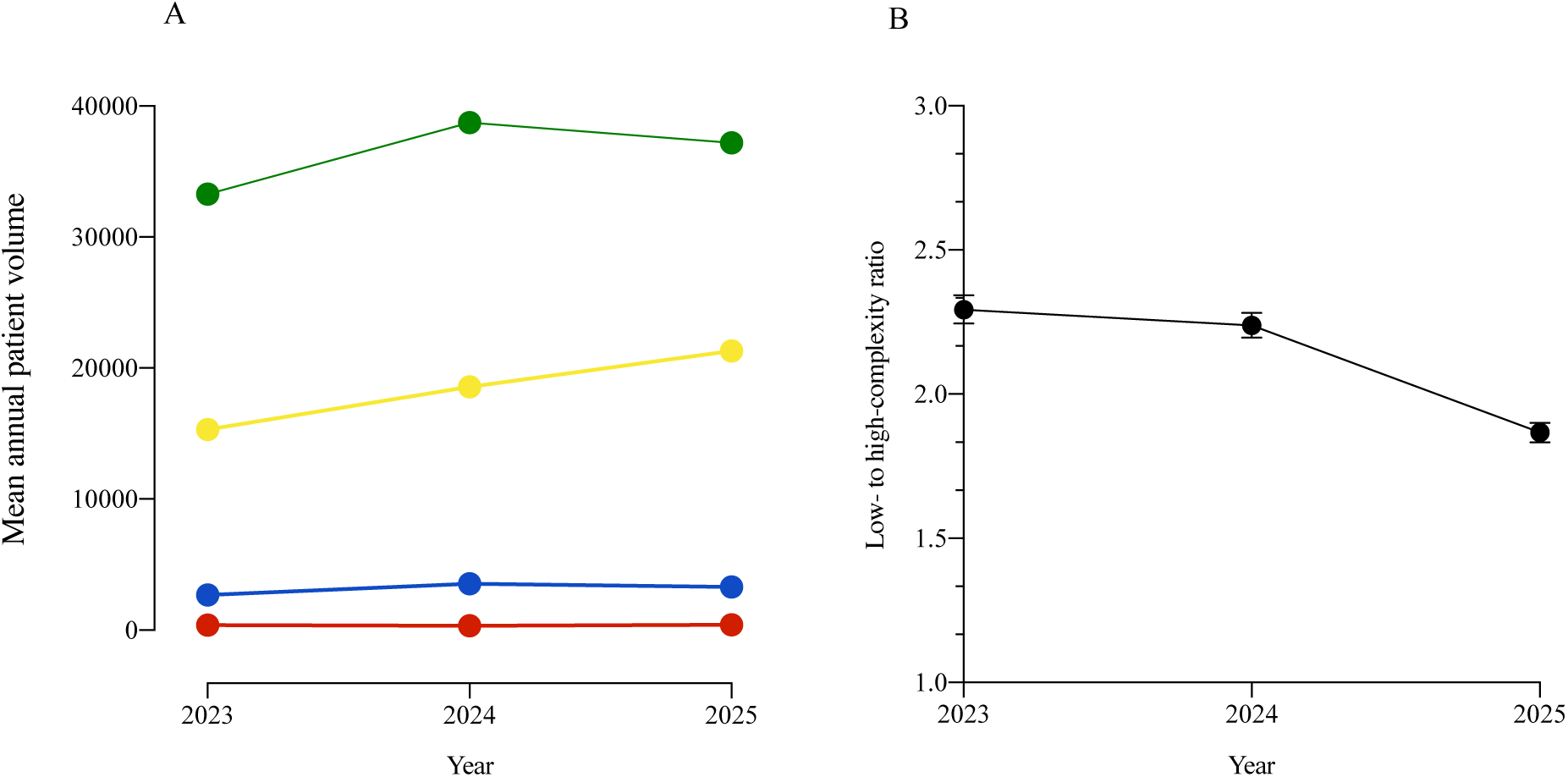
Distribution of visits according to risk classification and ratio between low and high complexity demand in the emergency sector of Santa Casa de Misericordia, Barra Mansa, RJ (2023–2025). (A) Temporal evolution and distribution monthly of visits according to risk classification, with statistically significant differences between the categories and periods evaluated by the Tukey multiple comparisons test (*p < 0.0001*). (B) Ratio between low (blue and green) and high complexity (yellow and red) visits, where the points represent the monthly ratio and the vertical bars indicate the 95% confidence interval (95% CI). The temporal trend analysis by linear regression did not reveal a statistically significant difference over the period (*p* = *0.342*), with the consistent predominance of low complexity over high complexity visits. Source: Prepared by the authors based on the hospital’s attendance records.

### Incompleteness rate of attendance records according to persistence and by risk classification

From the moment the data already analyzed reveal that the low complexity profile massively predominates in visits to the emergency sector, we evaluated the negative impacts of this overload, and during the analyzed triennium, there were out of 175.004 admissions in the emergency sector a high rate of visits did not have the due completion of medical records, mainly in relation to the ICD-10 registration reaching 24,33% (n = 42.586), with values comparable to the sum of high complexity visits. Among these visits without the ICD-10 code record, the significant predominance of visits is concentrated in the same low-complexity visits (green category with n = 31.040 and blue category with n = 3.673) records, in relation to high-complexity visits (yellow category with n = 7.867 and red category with n = 6). This data demonstrates that the incompleteness of medical records may be directly related to the intense overload of low-complexity cases, significantly compromising the quality of service, as it can consequently impact the generation of data and information for notification records and statistical flow in government databases. (Figure 4A).

**Figure 4.**
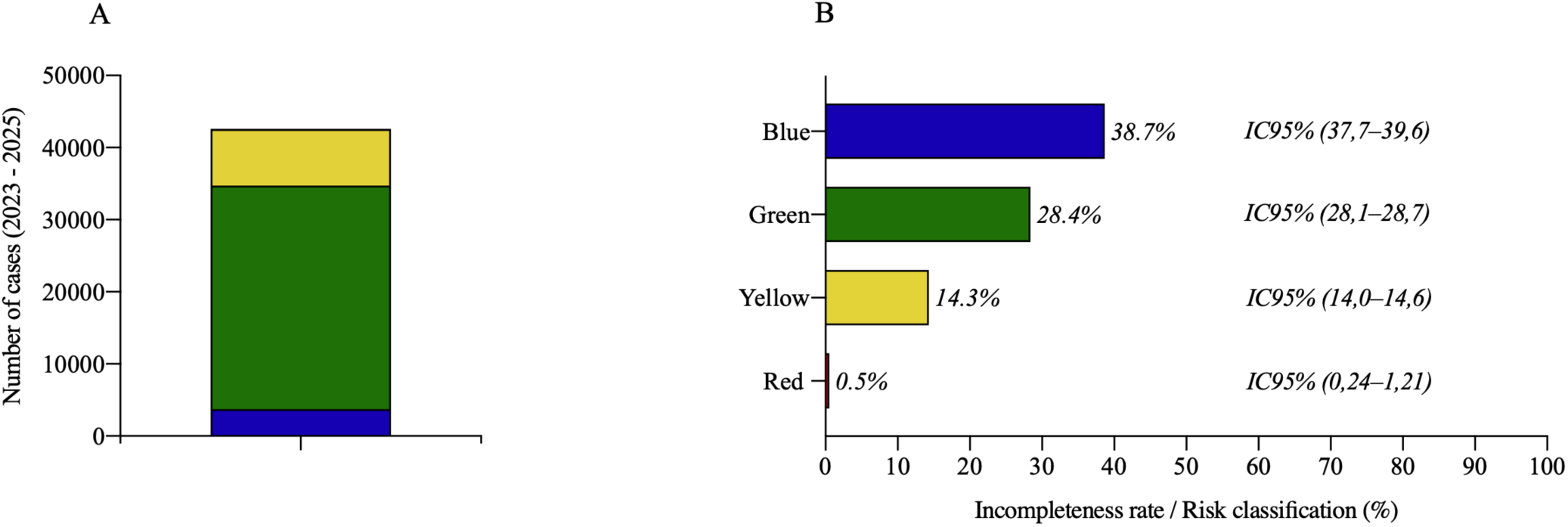
Cumulative distribution of emergency visits without ICD-10 code registration according to risk classification in the emergency department of Santa Casa de Misericordia, Barra Mansa, RJ (2023 – 2025; n = 42.586). (A) The stacked bar chart illustrates the total absolute volume by risk category (blue, green and yellow; red category is not seen in the chart due to its minute value compared to the other categories) in records lacking diagnostic coding. Chi-square test for proportions, showed a statistically significant difference in the distribution of frequencies between the risk categories in records without ICD-10 (*p < 0.0001*), confirming the expressive concentration of cases of lower complexity in this portion of medical records. (B) Proportion of incomplete records without ICD-10 code by risk category: Blue (38,7%; 95% CI: 37.7 – 39.6), Green (28,40%; 95% CI: 28.1 – 28.7), Yellow (14,30%; 95% CI: 14.0 – 14.6) and Red (0,5%; 95% CI: 0.24 – 1.21). Note: Values express percentages with their respective 95% confidence intervals (95% CI) Low complexity: 29,2% vs High: 14,00% -PR = 2.09 (95% CI: 2.05–2.12). Source: Prepared by the authors based on the hospital’s service files.

Proportional analysis revealed an incompleteness gradient inverse to severity the incompleteness rate was 38,70% (95% CI: 37.7 – 39.6) in the blue category, 28,4% (28,1 – 28,7) in the green,14,3% (14,0 – 14,6) in the yellow and0,5% (0,2 – 1,2) in the red. Visits of low complexity (green and blue) showed 29,20% incompleteness, against 14,00% in those of high complexity (yellow and red), configuring a prevalence ratio of 2.09 (95% CI: 2.05 – 2.12) (Figure 4B).

### Spatial Distribution and Territorial Organization of the Network

The list of all locations of the UBS in the municipality of Barra Mansa, RJ, was tabulated according to the unit name, Neighborhood, and distance from Santa Casa de Misericordia (Supplementary Table 2). The spatial analysis of the Primary Care network in Barra Mansa, RJ (n = 52), showed an asymmetric territorial arrangement in relation to Santa Casa de Misericórdia, as illustrated (Figure 5A), the Euclidean distances ranged from 0.75 km to 19.13 km, with a mean of 4.03 km and a standard deviation of ~ 3.56 km. The result was confirmed through the care coverage density map (Figure 5B) which demonstrates a strong spatial concentration of health establishments in the immediate surroundings of the hospital, revealing a decreasing density gradient towards the peripheral areas. The frequency histogram (Figure 5C) demonstrates, through linear correlation analysis between distance and unit frequency, a strong and statistically significant negative association (r = -0.7120; *p < 0.0001*; N = 52). The coefficient of determination (*R_2_ = 0,5069*) indicated that about 50.70% of the variation in the distribution of units is explained by the distance in relation to the reference hospital unit, confirming the network concentration gradient in the immediate surroundings and the progressive thinning in the more distant ranges.

**Figure 5.**
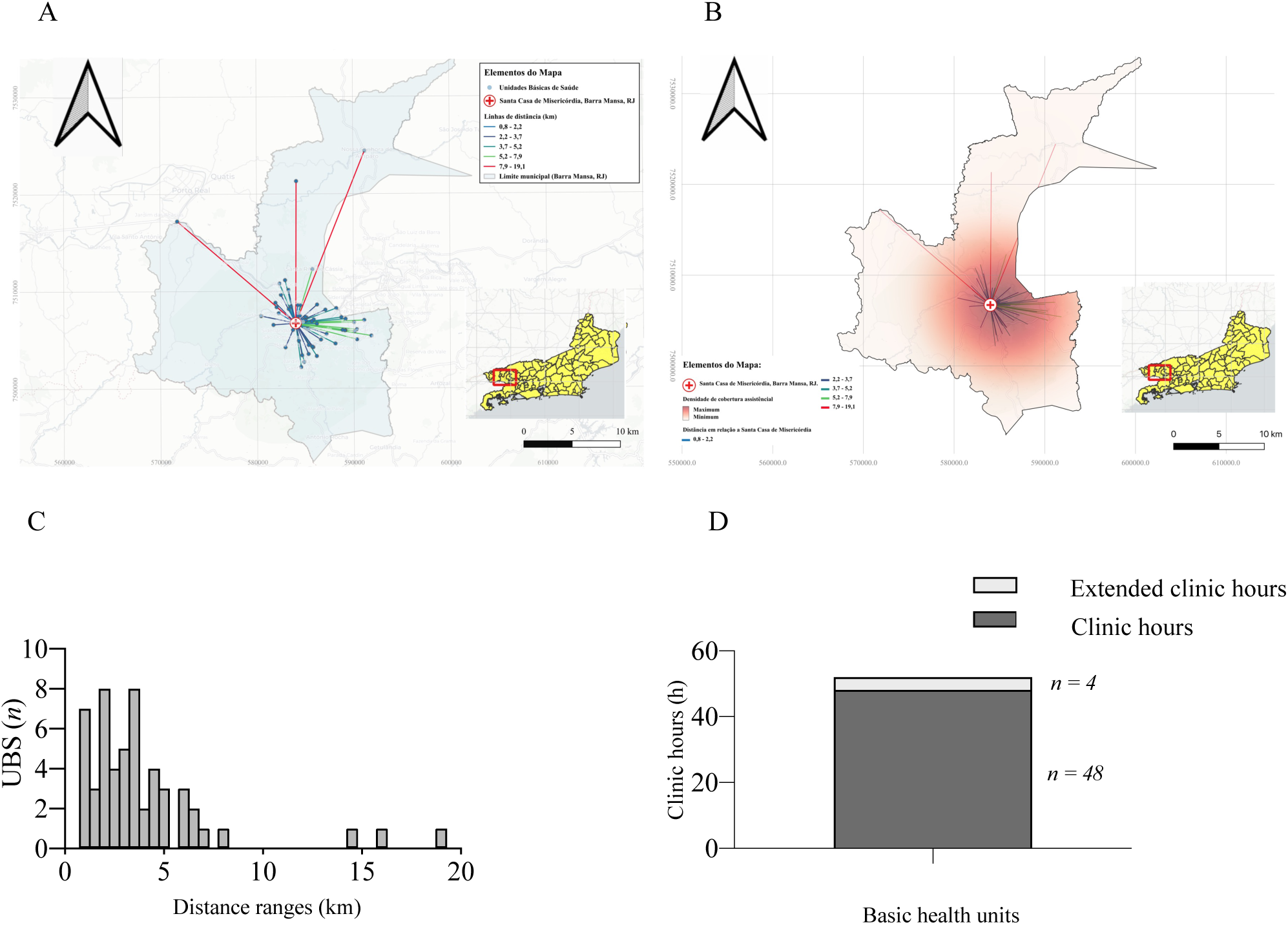
Spatial analysis and distribution of Euclidean distances between Basic Health Units (UBS) and the reference hospital (Santa Casa de Misericórdia) in Barra Mansa, RJ (n = 52). (A) Detailed distribution map of the frequency of UBS by distance ranges in kilometers. (B) Kernel density map evaluating the density of care coverage and linear distance lines. (C) Frequency histogram of distances in kilometers (mean = 4.03 km; SD = 3.56 km; minimum = 0.75 km; maximum = 19.13 km) (D) Comparative profile of UBS operating hours (extended hours, n = 4; daytime hours, n = 48). Note: Cartographic and geographic data obtained from official IBGE grids and records from the National Registry of Health Establishments (CNES/DATASUS). Source: Prepared by the authors (2026).

Furthermore, regarding the operational and temporal dimension of care, it was observed a significant restriction in coverage outside business hours, where only 4 units operate under an extended hours regime (Figure 5D), which contrasts expressively with the contingent of 48 units with exclusively daytime operation.

### Theoretical-explanatory chain model of the determinants of saturation in the tertiary hospital service (2023–2025)

The findings described in the present work allowed for the development of an explanatory model for the phenomenon of overcrowding in the analyzed emergency service. The model synthesizes the determination relationships that connect potential structural factors of origin with the saturation of tertiary care to its cascading impacts, highlighting that high demand may be related to a negative gradient of UBS concentration at the hospital unit and the temporal rigidity of care with only 4 units with extended operating hours, culminating in a hospital sector overloaded by more than 67.85% of low-complexity cases, and this problem implying serious informational problems through the critical increase in incomplete record rates of 24.33% (Figure S2).

## DISCUSSION

The present study demonstrates that the saturation of emergency services is not an isolated phenomenon, but the sequential effect of structural failures in the HCS, characterized by the inversion of the care flow, where PHC is unable to perform its function as the center coordinating care^1,2^. This problem was observed in the present work, since of the significant total volume of visits accumulated in the triennium from 2023 to 2025 (175,004), the largest portion was composed of low-complexity cases (67.85%), which corroborates the so-called “inverted pyramid” effect in the SUS, signaling that the operational rigidity of the basic network, which may be related to multiple factors such as limited operating hours and difficulties in access, directs the user to a more convenient option for accessing the public health system, the emergency sector^15,16^.

From a social perspective, the hospital has consolidated itself as a hub of safety immediate diagnostic and therapeutic, in addition to having, for the most part, a geolocation central within large urban centers^17^, and as illustrated by the explanatory model of figure S2, this hospital centrality is driven by greater ease of access and by the fragility of PHC in absorbing spontaneous demand during alternative hours. However, this dynamic disproportionately burdens the tertiary level, overloads care teams and compromises patient safety, forcing more serious cases to compete with outpatient demands for time and resource^15,16,18^.

Furthermore, from the perspective of spatial analysis and georeferencing, the data reveal that this overload may be being intensified by the very territorial layout of the health network, as the study of the distribution of the 52 health units in the municipality of Barra Mansa demonstrated average Euclidean distances of 4.03 km (ranging from 0.75 km to 19.13 km) in relation to the Santa Casa de Misericordia. The Kernel density map and linear correlation proved a strongly negative gradient of unit concentration in the immediate surroundings of the hospital, associated with a progressive emptying in the more distant bands.

Additionally, the temporal dimension aggravated this scenario, as only 4 units operated under an extended hours regime, compared to 48 units operating exclusively during the day, demonstrating that this hospital centrality added to the care void during alternative hours act by diverting spontaneous demand, channeling flows that should be absorbed by the basic network directly to the tertiary emergency room^4,19^.

All these findings point to several critical factors that together seem to feed back into each other, further intensifying the fragility of primary care and the overload of the tertiary health sector^20^. Among these damages caused by this disproportion, one of the most critical findings of this study is the relationship between care overload and the precariousness of health records, with a significant rate of incompleteness, since out of the 175.004 records analyzed, an alarming incompleteness rate of 24,33% (n = 42.586) was observed due to the absence of the ICD-10 code, being a serious flaw in the systematic production of health information. The distribution of this informational gap did not occur randomly; the data presented in the work demonstrate that there was an expressive concentration of medical record incompleteness exactly in low-complexity care (with thousands of records in the green and blue categories). Additionally, proportion analyses revealed an inversely proportional relationship between complexity and record incompleteness, where we observed that the lower the severity of the care, the higher the rate of medical records with incomplete entries, configuring a prevalence ratio of (PR ~2,09), making it clear that the increase in low-complexity demand is 2 times more likely to result in incomplete medical records. This result supports the premise that overcrowding by less severe cases physically and mentally overburdens the care teams, generating an environment of high turnover and rushed filling of medical records. Operational overload degrades the work process at reception and triage, weakening the culture of qualified documentary recording in favor of immediatist and fragmented care^21–24^.

As a consequence, this high rate of incomplete medical records severely compromises the epidemiological surveillance chain and public management with the systematic loss of basic diagnostic coding and corrupts the accuracy of vital national databases such as the Hospital Information System (SIH/SUS) and notification systems, generating statistical distortions that make the real epidemiological profile an unknown for managers25,26. Deprived of reliable data, public administrators encounter significant barriers to planning evidence-based interventions, allocating resources equitably, or correcting regional supply inequalities^26^.

It is concluded, therefore, that addressing this problem is not restricted to the physical expansion of emergency beds, requiring primarily the restructuring of PHC through the real expansion of the operating hours of basic units, the qualification of the welcoming spontaneous demand and interoperable computerization, shielding the system against the loss of data essential to the sovereignty of health management.

## Data availability statement

The entire dataset supporting the results of this study was published in the article itself.

## Ethics committee approval statement

The research project was submitted to and approved by the Research Ethics Committee (CEP), under the Certificate of Presentation of Ethical Appreciation (CAAE) no. 92484025.4.0000.8044

## Artificial intelligence use statement

During the preparation of the manuscript, the authors used Artificial Intelligence only for refinement and spelling correction stages of the manuscript writing. The authors declare that they did not use artificial intelligence resources for data analysis nor for the construction of figures, tables, or graphs.

## Author contribution

Alexandre Rodrigues de Souza Soares Conceptualization: methodology, data collection, writing of the original version of the manuscript.

Tamyres Silva Salles Cardoso; Yago Aragão Lessa; Luís Gustavo Ferreira de Souza; João Victor Cordeiro da Silva; Rachel Rocha Ferreira: Visualization, data organization, data collection, data analysis, editing of the manuscript.

Aluana Santana Carlos and André Manoel Correira dos Santos: Data analysis, supervision of the review and editing of the manuscript.

André Costa Ferreira: Team coordination, conceptualization, methodology, supervision, review, data analysis, editing, and finalization of the manuscript.

## Supplementary material

**Supplementary table 1.**
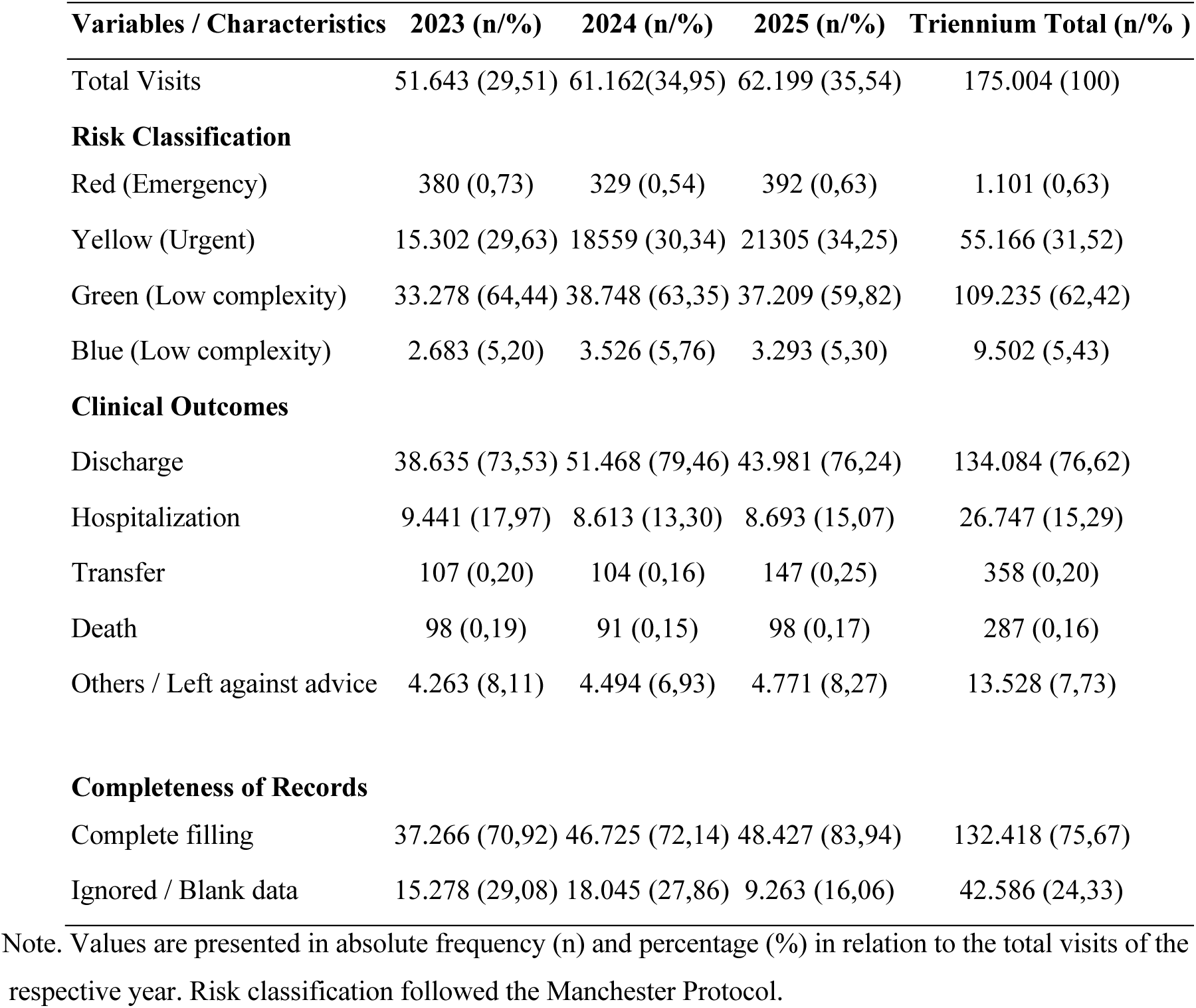
Distribution of emergency visits according to annual volume, risk classification by the Manchester protocol, clinical outcomes, and completeness of records in the emergency department of Santa Casa de Misericórdia, Barra Mansa, RJ, during the 2023-2025 triennium.

| **Variables / Characteristics** | **2023 (n/%)** | **2024 (n/%)** | **2025 (n/%)** | **Triennium Total (n/% )** |
|---|---|---|---|---|
| Total Visits | 51.643 (29,51) | 61.162(34,95) | 62.199 (35,54) | 175.004 (100) |
| **Risk Classification** |  |  |  |  |
| Red (Emergency) | 380 (0,73) | 329 (0,54) | 392 (0,63) | 1.101 (0,63) |
| Yellow (Urgent) | 15.302 (29,63) | 18559 (30,34) | 21305 (34,25) | 55.166 (31,52) |
| Green (Low complexity) | 33.278 (64,44) | 38.748 (63,35) | 37.209 (59,82) | 109.235 (62,42) |
| Blue (Low complexity) | 2.683 (5,20) | 3.526 (5,76) | 3.293 (5,30) | 9.502 (5,43) |
| **Clinical Outcomes** |  |  |  |  |
| Discharge | 38.635 (73,53) | 51.468 (79,46) | 43.981 (76,24) | 134.084 (76,62) |
| Hospitalization | 9.441 (17,97) | 8.613 (13,30) | 8.693 (15,07) | 26.747 (15,29) |
| Transfer | 107 (0,20) | 104 (0,16) | 147 (0,25) | 358 (0,20) |
| Death | 98 (0,19) | 91 (0,15) | 98 (0,17) | 287 (0,16) |
| Others / Left against advice | 4.263 (8,11) | 4.494 (6,93) | 4.771 (8,27) | 13.528 (7,73) |
| **Completeness of Records** |  |  |  |  |
| Complete filling | 37.266 (70,92) | 46.725 (72,14) | 48.427 (83,94) | 132.418 (75,67) |
| Ignored / Blank data | 15.278 (29,08) | 18.045 (27,86) | 9.263 (16,06) | 42.586 (24,33) |

Note. Values are presented in absolute frequency (n) and percentage (%) in relation to the total visits of the respective year. Risk classification followed the Manchester Protocol.

**Figure S1.**
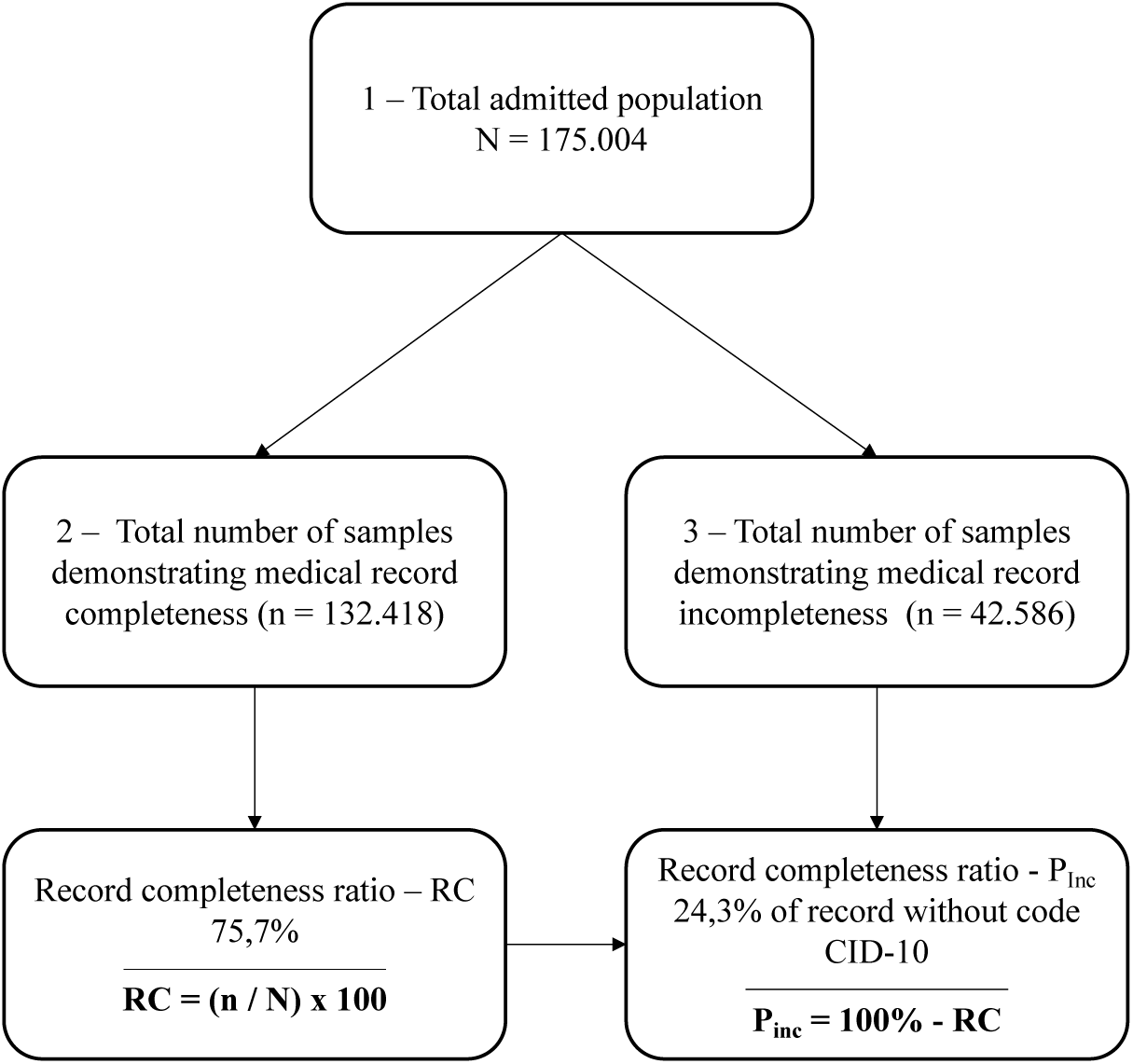
Sample selection flowchart based on international STROBE guidelines. Flowchart of the sample selection (STROBE) and indicators of completeness and incompleteness of medical records in the hospital emergency service of Barra Mansa, RJ (2023–2025). Source: Prepared by the authors (2026).

**Supplementary Table 2.**
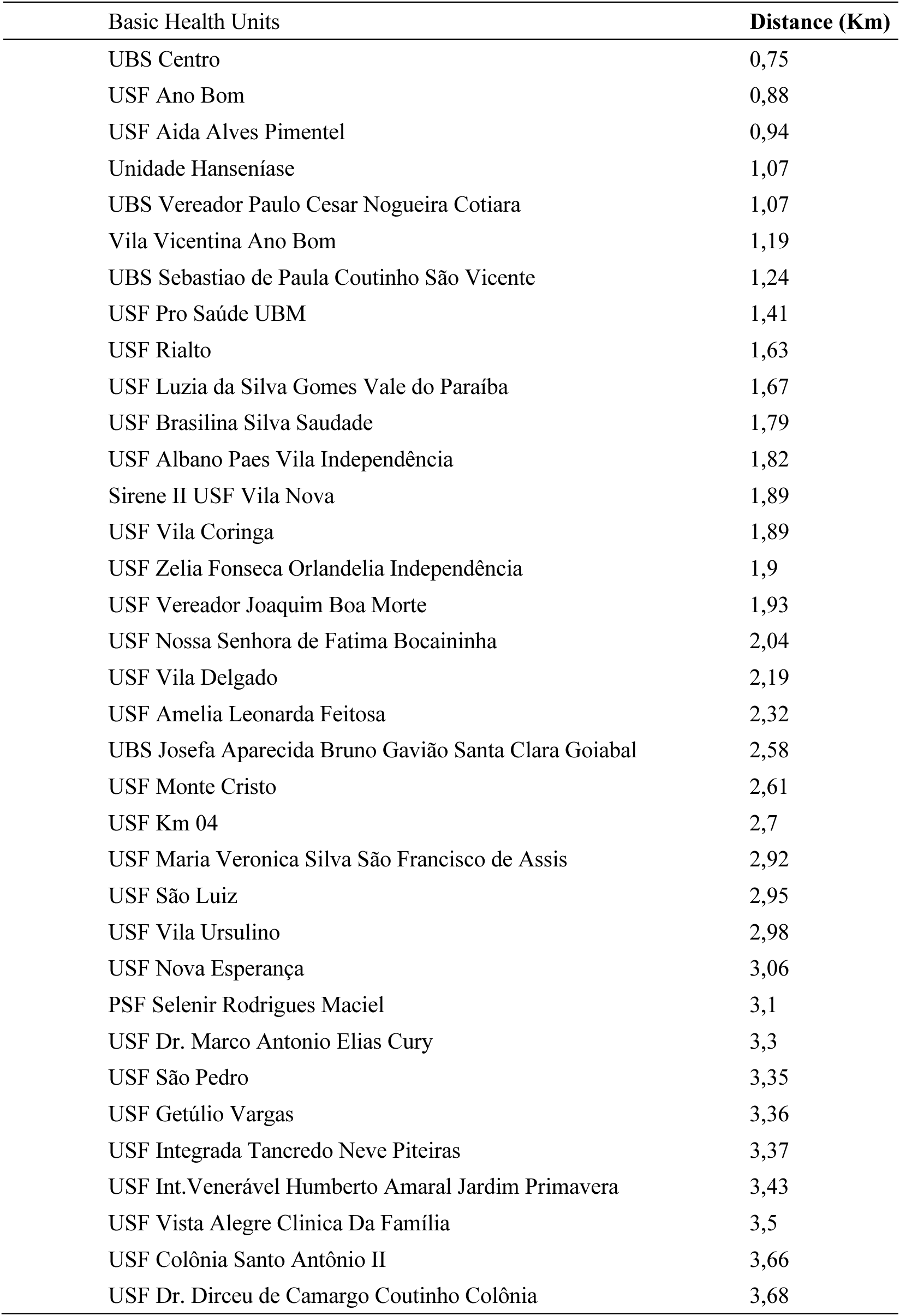
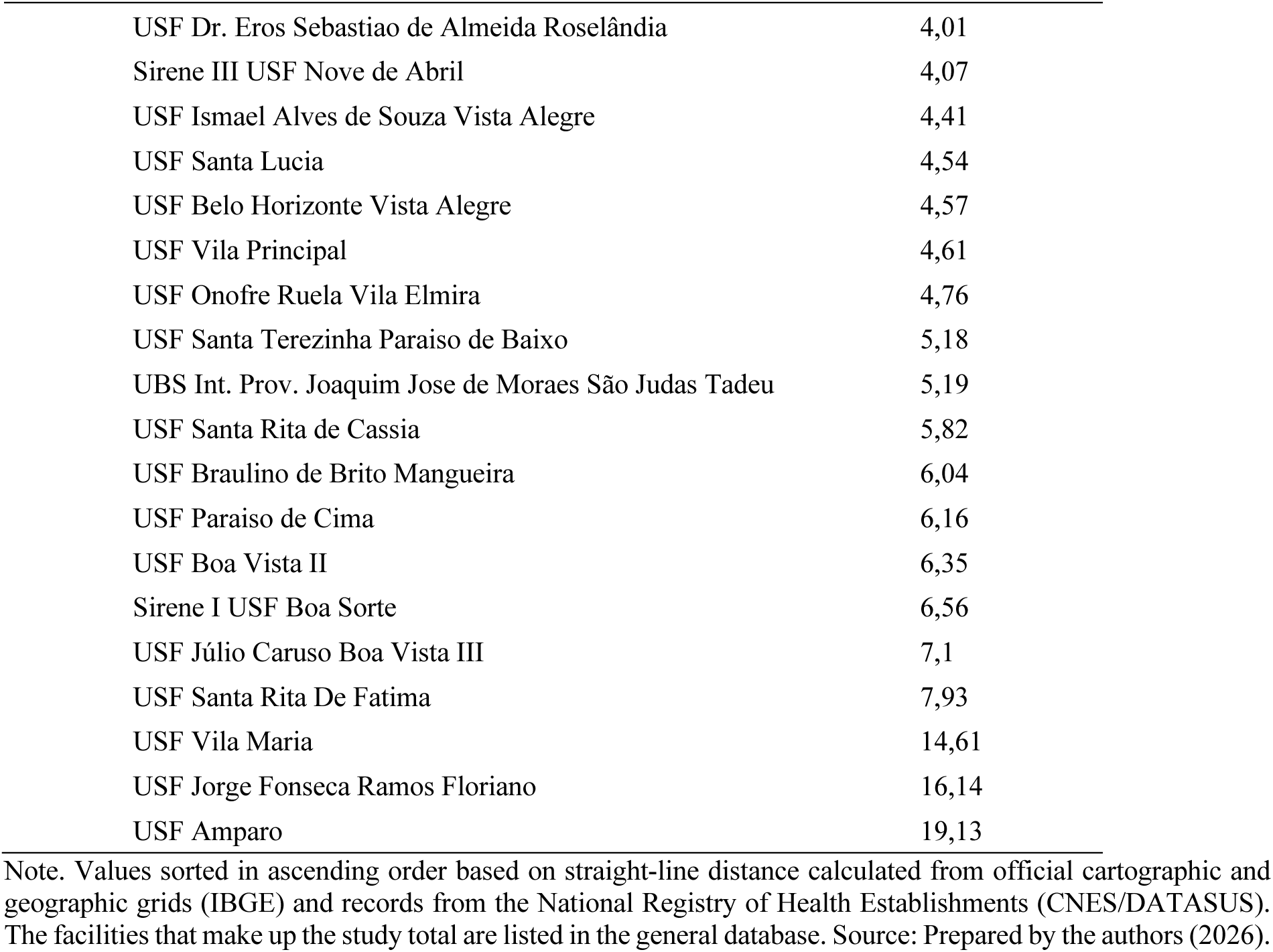
List of Basic Health Units (UBSs) and their respective Euclidean distances (straight-line in kilometers) to the reference hospital unit (Santa Casa de Misericordia), Barra Mansa, RJ.

**Figure S2.**
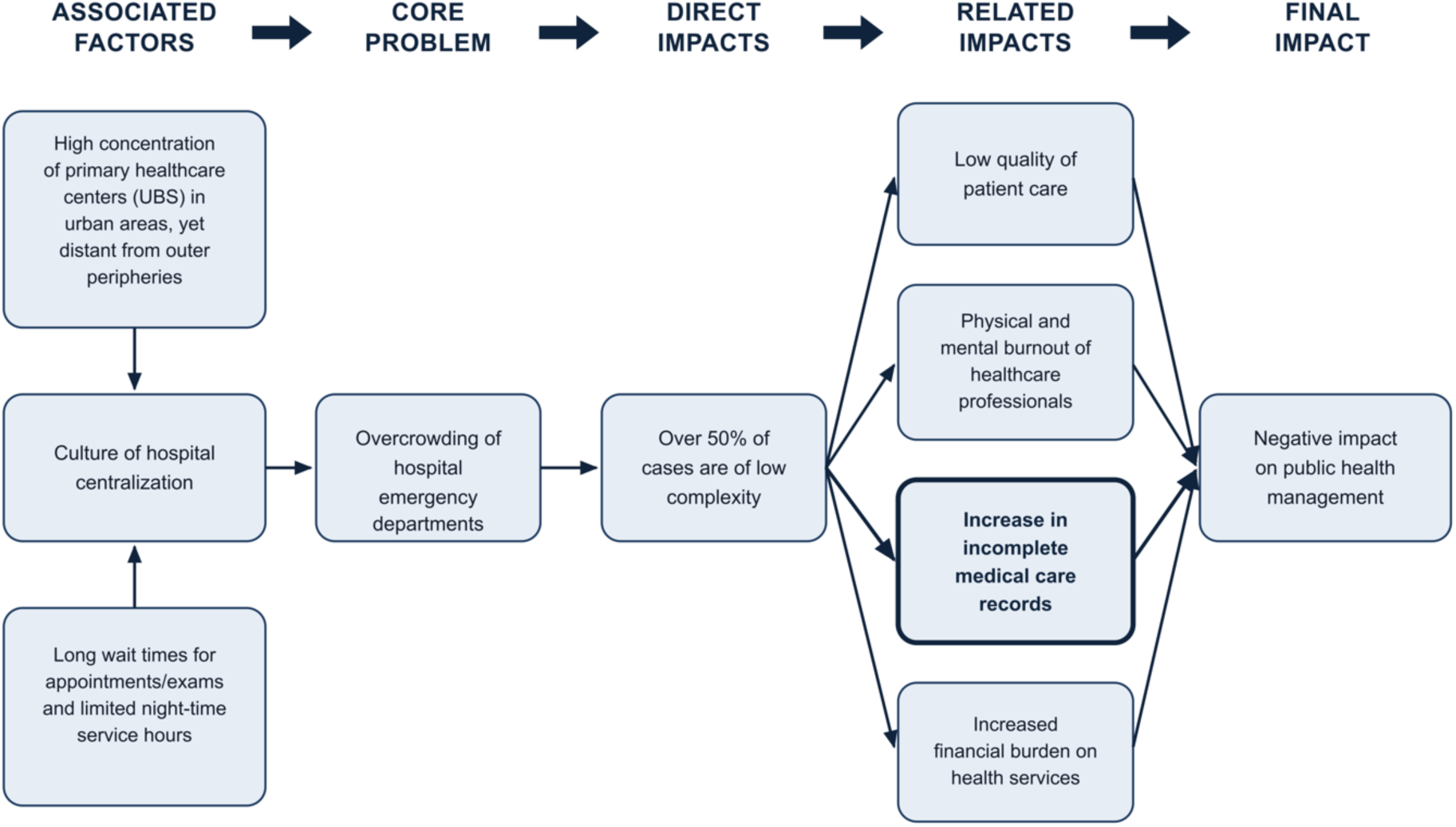
Theoretical-explanatory chain model of the determinants of saturation in hospital service tertiary (2023–2025). Chain flowchart illustrating the logical progression of overcrowding. Source: Prepared by the authors (2026), using data from Google Maps/IBGE and operational data from the study.

